# Phonemic awareness deficits in an alphasyllabary language: Effects of task type and linguistic complexity in children with dyslexia

**DOI:** 10.64898/2026.04.02.26349894

**Authors:** Anjana Soman, Sonu S. Dev, Rajith Ravindren

**Affiliations:** Department of Clinical Psychology, Institute for Communicative and Cognitive Neurosciences (ICCONS), Shoranur, Kerala, India; Department of Clinical Psychology, Institute of Mental Health and Neurosciences (IMHANS), Kozhikode, Kerala, India; Department of Psychiatry, Institute of Mental Health and Neurosciences (IMHANS), Kozhikode, Kerala, India

**Author notes:** Corresponding author Rajith Ravindren.

**Keywords:** dyslexia, phonemic awareness, pseudoword processing, alphasyllabary, Malayalam

## Abstract

Phonemic awareness deficits are a core feature of Specific Learning Disorder with impairment in reading (dyslexia). Examining how task- and language-specific factors influence these deficits in alphasyllabary languages may help clarify the cognitive mechanisms underlying reading impairment in dyslexia.

Thirty children with Specific Learning Disorder with impairment in reading (dyslexia) and 29 age-matched typically developing children were administered phoneme blending (words and pseudowords) and segmentation tasks in Malayalam. The effects of age and consonant clusters on task performance were examined.

Children with dyslexia performed significantly worse than typically developing controls across most phonemic awareness tasks, with the largest deficits observed in pseudoword and word blending, and smaller deficits in segmentation. No significant group difference was observed for blending after initial phoneme deletion. In typically developing children, age showed strong positive correlations with phonemic awareness performance across most tasks, whereas the dyslexia group showed weak or absent correlations, except in word blending and initial phoneme deletion. Consonant clusters significantly affected performance in both groups, with more pronounced deficits in the dyslexia group.

Phonemic awareness deficits in dyslexia within alphasyllabary languages such as Malayalam are more prominent in tasks with limited lexical support, such as pseudoword blending. These deficits vary across task demands and linguistic complexity. While phonemic awareness improves with age in typically developing children, the trajectory of improvement is less consistent in children with dyslexia. These findings suggest that phonemic awareness deficits are a core feature of dyslexia across languages, while orthographic and linguistic characteristics of writing systems influence their manifestation.

## Introduction

Specific Learning Disorder with impairment in reading (dyslexia) is a common neurodevelopmental condition with a prevalence rate of 4%-9% (American Psychiatric Association, 2013). It is characterized by persistent difficulties in reading accuracy, fluency, and comprehension, which result in poor academic performance, poor self-esteem, and increased risk of anxiety and depressive symptoms. Co-morbid conditions like ADHD and conduct disorder further worsen the long-term outcome of this disorder (Moll et al., 2014).

Earlier research on the neurobiology of dyslexia has suggested impairments in the magnocellular pathway, cerebellar defects, and temporal processing defects as the underlying pathology (Ziegler & Goswami, 2005). However, converging evidence across multiple languages has validated phonological processing, particularly phonemic awareness, as the primary impairment in this disorder. Phonological awareness proceeds along a continuum, from sensitivity to larger units of sound (syllables, onsets, and rimes) to awareness of smaller units, namely phonemes (Goswami, 2010).

Phonemic awareness is the ability to identify, analyse, and manipulate individual speech sounds and is widely recognised as a foundational skill for reading acquisition and a core deficit in children with reading disorders (Stanovich, 1988; Melby-Lervåg, 2012). While the relationship between phonemic awareness and dyslexia has been extensively studied in alphabetic languages such as English, there is comparatively limited research examining this relationship in languages with shallow orthographies and alphasyllabary writing systems.

This gap is particularly relevant in the Indian context, where languages such as Malayalam exhibit distinct phonological and orthographic characteristics that may influence how reading disorders manifest (Nag & Snowling, 2012).

Malayalam is an alphasyllabary language derived from the Brahmi script in which phonological information is primarily encoded at the syllabic (Akshara) level (Haridas et al., 2024). Akshara units typically consist of consonants followed by vowels as diacritics. Although the transparent orthography of Malayalam facilitates decoding, its morphological structure is complex due to its agglutinative nature and the presence of multiple ligature rules (Haridas et al., 2024. Besides, the vowel diacritics may appear in multiple spatial positions (left, right, or above consonants). The large number of symbols and the complexity of the orthography increase the cognitive demand of learning to read. Consequently, reading difficulties in dyslexia within the Akshara system may arise not only from phonemic deficits but also from orthographic complexity, as in alphabetic languages (Nag, 2007).

Research on Indic scripts suggests that children learning to read in such systems develop syllable-level awareness earlier than phoneme-level awareness, as orthographic units map more directly onto syllables than individual phonemes. (Nag & Snowling, 2012). But studies examining the role of phonemic awareness in alphasyllabary reading have given inconsistent findings (Somashekara et al., 2014). Most of these studies were conducted on typically developing children, and the role of phonemic awareness deficits in alphasyllabary language-speaking children with dyslexia remains underexplored. The relative contributions of task type (blending vs segmentation), linguistic complexity (consonant clusters), and developmental factors (age) are not well understood.

The study aims to examine phonemic awareness deficits in Malayalam-speaking children with dyslexia with a focus on the influence of task type, age, and linguistic factors on performance. This study seeks to answer whether phonemic awareness is a universal feature of dyslexia or if its manifestation is skewed by the orthographic and linguistic features of alphasyllabary languages. Answering these questions will help in developing linguistically appropriate assessment tools and targeted interventions for this disorder in Indian languages.

## Methodology

### Study Design

The study employed a descriptive, cross-sectional comparative research design to examine differences in phonemic awareness between children diagnosed with Specific Learning Disorder with impairment in reading (dyslexia) in Malayalam and typically developing peers.

Thirty children aged 8 to 13 years, diagnosed with specific learning disorder with impairment in reading as per DSM-5 criteria, and with an IQ above 86, were included in the experimental group. The children were native Malayalam speakers. The dyslexia diagnosis was confirmed using the NIMHANS Index for Specific Learning Disabilities (Kapur et al., 1991). Malin’s Intelligence Scale for Indian Children (MISIC) was used for IQ assessment (Malin, 1971). Children with hearing impairment, comorbid neurodevelopmental (e.g., autism) or psychiatric disorders (e.g., depression), speech disorders, or prior remedial intervention were excluded. Children with comorbid attention deficit hyperactivity disorder (ADHD) were excluded to minimise the effects of inattention in the phonemic awareness tasks. The DSM-5-TR Parent/Guardian-Rated Level 1 Cross-Cutting Symptom Measure for psychiatric screening was used to screen for psychiatric symptoms (American Psychiatric Association, 2022). The experimental group was drawn from a tertiary care clinical setting. The age range was selected because phonemic awareness skills are sufficiently developed at this stage to allow reliable assessment and comparison.

For the control group, 29 typically developing children, aged 8 to 13 years, were recruited from schools in the Ernakulam district. The control group included children with average or above-average IQ with no history of dyslexia, speech disorders or psychiatric disorders. NIMHANS Index for Specific Learning Disabilities, Malin’s Intelligence Scale for Indian Children and DSM-5-TR Parent/Guardian-Rated Level 1 Cross-Cutting Symptom Measure for psychiatric screening were used in the selection of the children in the control group.

Efforts were made to ensure comparability between groups in terms of age, schooling, and socioeconomic background.

### Measures

1. **Sociodemographic Data Sheet**: A researcher-developed form used to collect information on age, gender, class, medium of instruction, and clinical profile.
2. **Phonemic Awareness Word List (Malayalam)**: A structured assessment tool was developed to evaluate phoneme blending, segmentation, and deletion. The phonemic awareness word list was developed based on established paradigms used in prior phonemic awareness research and adapted to the Akshara structure of Malayalam.

Though formal large-scale psychometric validation of the word list was not conducted, content validity was established through expert review, and pilot testing was undertaken to ensure item clarity and difficulty. The list included words beginning with vowels and consonants, consonant clusters, words with increasing phonemic complexity, and a set of meaningless pseudowords to assess phonemic processing independent of lexical knowledge.

#### Blending - Pseudowords

The words were made by changing letters of common Malayalam words. A list of 10 words was used. The list included five words with five phonemes, two words with six phonemes, and one word each with 7,8,9 phonemes. Six words had one consonant cluster, while one word had two consonant clusters.

#### Blending - Words

Fourteen common words used in the school textbooks were used-Four words with six phonemes, three words with five phonemes, two words each with 3,4,8 phonemes, and one word with seven phonemes were used. Four words had one consonant cluster, while one word had two consonant clusters.

#### Segmenting Words

Fourteen common words used in school textbooks were used. Five words with seven phonemes, four words with six phonemes, two words each with 3 and 4 phonemes, and one word with 8 phonemes were used. Four words had consonant clusters.

#### Blending after deleting the initial and final phonemes

Five words each were used for blending after deleting the first and last phoneme, respectively.

### Procedure

Following participant selection, written informed consent was obtained from parents, and assent was obtained from the children. The experimental group was assessed at a clinical setting, while the control group was assessed in school settings after obtaining institutional permission.

All the participants underwent hearing screening to rule out auditory impairment. IQ assessment, psychiatric screening, and confirmation of dyslexia diagnosis were completed before phonemic awareness testing.

The phonemic awareness word list was pre-recorded and presented binaurally through standardized headphones at approximately 65 dB SPL, with uniform volume settings across participants. Practice trials were administered before testing. The assessment included tasks of phoneme blending, segmentation, blending of meaningless words, and phoneme deletion (initial and final sounds). Responses were recorded and scored using a standardized scoring protocol based on correct phonemes. A phoneme-level scoring approach was used to capture partial performance, as binary word-level scoring may underestimate phonemic processing ability.

For blending, a score of 1 was given if the students were able to blend a consonant and a vowel or two consonants. The total score for each word was then calculated.

For segmenting, a score of 1 was given for each phoneme, and the total score was calculated for each word.

For blending after deleting initial and final phonemes, a score of 1 was given for the correct response. A total phoneme score was calculated by adding the scores of the five tests.

### Ethical Clearance

The study was approved by the institutional ethics committee of the host institute. Written informed consent from parents and assent from children were obtained before participation. Confidentiality of the data was maintained, participation was voluntary, and children were allowed to withdraw at any stage. The study involved no physical or psychological harm to the participants.

### Statistical Analysis

Data were analysed using JASP statistical software. Descriptive statistics (mean and standard deviation) were computed for phonemic awareness scores. Normality assumptions were examined before inferential analysis. As the data were not normally distributed, non-parametric tests were used for comparison. Mann-Whitney U tests were conducted to compare phonemic awareness performance between the experimental and control groups. For subgroup analysis within the group, the Wilcoxon signed-rank tests were used. Effect sizes (rank biserial correlation) were calculated to estimate the magnitude of group differences. The level of statistical significance was set at p < 0.05. Correlation tests (Spearman’s) were used to find the association between age and phonemic score. To examine whether group differences in phonemic awareness were independent of age, a linear regression analysis was conducted as regression is robust to mild deviations from normality. The phonemic awareness score was entered as the dependent variable, with group (0 = dyslexia, 1 = control) and age as predictors. This approach allowed us to assess whether group differences persisted after controlling for developmental effects.

## Results

### A. Participant characteristics

The dyslexia (*n* = 30) and control (*n* = 29) groups were comparable in age, class level, and gender distribution. The mean age of the dyslexia group was 11.43 years (SD = 1.46), while the control group had a mean age of 11.96 years (SD = 1.72). The case group had 5 girls and 25 boys, while the control group had 6 girls and 23 boys. All the participants were right-handed.

### B. Phonemic awareness accuracy

We compared cases and controls using the Mann-Whitney U test for differences in performance in the phonemic awareness tasks. Children with dyslexia performed significantly worse than controls across most phonemic awareness tasks, with the largest deficits observed in pseudoword blending and word blending, and smaller deficits in segmentation. No significant difference was observed for blending after initial phoneme deletion. (**Table 1**)

**Table 1:** Comparison of phonemic awareness tasks between children with dyslexia(cases) and typically developing children(controls)

|  | **U** | **P value** | **Effect size (Rank-Biserial Correlation)** |
| --- | --- | --- | --- |
| **Blending pseudowords** | 93.500 | < .001 | 0.785 |
| **Blending words** | 114.000 | < .001 | 0.738 |
| **Segmenting words** | 293.500 | 0.030 | 0.325 |
| **Blending after deleting the initial phoneme** | 361.500 | 0.235 | 0.169 |
| **Blending after deleting the final phoneme** | 130.500 | < .001 | 0.700 |
| **Total Phoneme score** | 104.000 | < .001 | 0.761 |

\* $p < .05$ indicates statistical significance.
U=Mann-Whitney U statistic

### C. Age and phonemic awareness tasks

Spearman’s correlation tests were used to determine the association between age and phonemic awareness tasks. In typically developing children, age showed strong positive correlations with phonemic performance across most tasks (ρ = 0.70–0.78, p < .001). In contrast, the dyslexia group showed weak or absent correlations for most tasks except in word blending (ρ = 0.38) and initial phoneme deletion (ρ = 0.61) (**Table 2)**

**Table 2:** Correlation of age with phonemic awareness tasks in cases and typically developing children(controls)

| **Task** | **Cases rho** | **Cases P value** | **Controls rho** | **Controls P value** |
| --- | --- | --- | --- | --- |
| Blending pseudowords | 0.03 | 0.87 | 0.70 | **<0.001** |
| Blending words | 0.38 | **<0.001** | 0.78 | **<0.001** |
| Segmenting words | 0.24 | 0.20 | 0.70 | **<0.001** |
| Blending after deleting the initial phoneme | 0.61 | **<0.001** | 0.10 | 0.61 |
| Blending after deleting the final phoneme | 0.35 | 0.06 | 0.42 | **0.002** |
| Total phoneme score | 0.25 | **0.18** | 0.74 | **<0.001** |

Rho- $\rho$ = Spearman's rank correlation coefficient.

### D. Phonemic awareness in words with consonant clusters

We measured how performance was affected in words with consonant clusters. We used Wilcoxon-signed rank to compare the difference in performance in both cases and controls.

Consonant clusters significantly affected performance in both groups, but the magnitude and pattern of this effect differed. In children with dyslexia, consonant clusters produced large performance decrements across tasks. In contrast, typically developing children showed a smaller effect size in blending words, suggesting more efficient processing or compensatory strategies. (**Table 3)**

**Table 3.** Performance on phonemic awareness tasks with and without consonant clusters in children with dyslexia(cases) and typically developing children(controls)

| **Task** | **Cases:**<br>**With**<br>**Cluster**<br>**Mean (SD)** | **Cases:**<br>**Without**<br>**Cluster**<br>**Mean (SD)** | **Cases p-**<br>**value**<br>**(effect**<br>**size)** | **Controls:**<br>**With Cluster**<br>**Mean (SD)** | **Controls:**<br>**Without**<br>**Cluster Mean**<br>**(SD)** | **Controls p-**<br>**value**<br>**(effect size)** |
| --- | --- | --- | --- | --- | --- | --- |
| Blending<br>pseudowords | 0.25 (0.34) | 0.68 (0.93) | **0.017**<br>**(0.61)** | 2.04 (0.98) | 2.40 (1.01) | **0.006 (0.61)** |
| Blending<br>words | 0.45 (0.62) | 1.03 (0.72) | **<0.001**<br>**(0.90)** | 2.22 (1.10) | 2.15 (0.83) | **0.032 (0.45)** |
| Segmenting<br>words | 0.10 (0.31) | 0.50 (0.35) | **<0.001**<br>**(0.80)** | 3.99 (3.35) | 3.22 (2.64) | **<0.001**<br>**(0.92)** |

Values are presented as mean (standard deviation). P values were derived using the Wilcoxon signed-rank test comparing performance on stimuli with and without consonant clusters within each group.

### E. Regression analysis

Linear regression analysis was conducted to assess whether group differences in phonemic awareness persisted after controlling for age. The overall model was significant, F (2,56) = 54.16, p < .001, explaining 65.9% of the variance in phonemic awareness scores (R^2^ = .659).

Both group and age emerged as significant predictors. Group was a strong predictor (B = 71.14, SE = 9.02, t = 7.89, p < .001), indicating that controls had significantly higher phonemic awareness scores than children with dyslexia. Age was also a significant predictor (B = 15.28, SE = 2.84, t = 5.37, p < .001), suggesting that phonemic awareness increased with age.

The group differences remained significant after controlling for age, indicating that the observed deficits in phonemic awareness in the dyslexia group were not attributable to age differences.

## Discussion

This study demonstrates that children with dyslexia have deficits in phonemic awareness tasks even in an orthographically transparent alphasyllabary language. The persistence of group differences after controlling for age supports a disorder-specific phonemic awareness deficit rather than a simple developmental delay. This is consistent with the evidence that phonemic awareness deficits are a core and potentially universal feature of dyslexia (Ziegler et al., 2010; Tang et al., 2025). Our findings challenge the assumption that phoneme-level deficits are less relevant in dyslexia in syllable-based scripts.

Previous research has consistently shown that phonemic awareness deficit is a core feature of dyslexia in alphabetic languages, with more severity observed in orthographically deep languages like English, French, and Portuguese. Studies have found that children learning English show poor word and non-word reading performance after grade 1 as compared to other transparent European languages, reflecting inconsistent grapheme-phoneme mapping in English (Seymour et al., 2003; Frith et al., 1998). Similar results were also observed in French (Porpodas, 1999).

However, deficits in phonemic awareness in dyslexia are not confined to alphabets with deep orthographies. Cross-linguistic studies have proven that phonemic awareness tasks are deficient in transparent alphabetic languages like Greek and Dutch, though with lesser severity (Porpodas, 1999; De Jong & van der Leij, 2003). These findings support the view that serial processing demands during phoneme blending may burden the cognitive resources for text understanding, resulting in reading difficulties in dyslexia (Ziegler et al., 2003).

In alphasyllabary languages, though print is represented in syllables, phoneme markers are also important. The relative contribution of syllabic and phonemic awareness in reading problems is debated. While some authors have found that syllable awareness is more important, recent studies have shown that phonological awareness is also critical in reading in alphasyllabary languages. These studies focused on typically developing children. The present study addresses this gap by specifically focusing on clinically defined children with dyslexia and examining their phonemic awareness deficits.

A key finding of this study is the disproportionate impairment in pseudoword blending, showing the largest effect size. Pseudoword tasks specifically check phonemic awareness by removing contextual and lexical cues. The poor performance in pseudowords blending compared to real words shows that children with dyslexia rely more on lexical support for word reading. This may reflect word superiority effects as proposed in the interactive activation model (Rumelhart & McClelland, 1982). Top-down effects from known words may help in blending in case of real words, but their absence in pseudowords exposes the underlying deficits in phonemic awareness.

Blending is considered more important in reading, especially in Malayalam, as the rules of ligature called sandhi are important in word formation (Haridas et al., 2024). Although blending showed the greatest impairment, we observed significant deficits in word segmenting also. This is clinically relevant as most of the children with dyslexia also have difficulties in spelling and writing. In contrast, we found that children did not differ in blending after deleting initial phonemes. It has been suggested that the initial phoneme awareness is a more important predictor of reading abilities in less skilled readers, though our findings are contrary to this view (Hulme et al., 2002). One possible explanation is that the initial phoneme deletion tasks do not check phoneme awareness, as the orthographic image may aid in performing these tasks (Goswami, 2002). The initial phoneme identification may be aided by syllable-level representation in the alphasyllabary, making it less prone to phoneme-level task deficits. This may also explain our findings. Or these tasks might have reached the ceiling levels in this age group. However, for final phoneme deletion, children with dyslexia did exhibit deficits, indicating that phoneme manipulation tasks are differentially affected in dyslexia.

We found that age was positively associated with performance in all the phonemic awareness tasks in the control population except the initial deletion. This is in line with the phonological grain size theory, which states that children first gain awareness of syllables, then of onset–rime units, and then phonemes (Goswami, 2010). It has been argued that the knowledge of phonemes does not occur naturally - it develops only if children are taught literacy (Goswami, 2002). Our finding of increasing phonemic awareness task scores with age in typically developing children supports this theory. In Malayalam, where Akshara represent syllabic knowledge, this finding becomes even more important. The lack of change in initial phoneme deletion with age in typically developing children may be due to the ceiling effect, as initial phoneme knowledge is easily learned in a syllabic system (Nag & Snowling, 2012; Goswami, 2002).

In contrast, children with dyslexia showed age-related change only in blending normal words and blending words after deleting initial phonemes. The word blending may be aided by the word superiority effect (Rumelhart & McClelland, 1982), while the initial phoneme effect might be due to the syllable knowledge of the language. This indicates that the phonemic awareness development in dyslexia is uneven and task-dependent, rather than uniformly delayed. Certain phonemic tasks improve with age -possibly through the language-specific compensatory mechanisms, while others remain persistently impaired.

We also evaluated how the linguistic complexity is affected in typically developing children and those with dyslexia. We examined the effect of consonant clusters, which are especially found in an agglutinative language like Malayalam (Haridas et al., 2024). We found that both cases and controls have difficulties in phonemic awareness tasks involving words with consonant clusters. But the severity of the deficits was less in control children for real words. This supports the view that dyslexia involves both phonemic deficits and reduced resilience to linguistic complexity.

Our study has several limitations. The use of chronologically age-matched controls rather than reading age-matched controls would have strengthened the interpretations of our study. The relatively small sample size and the use of a newly developed phonemic awareness tool without established psychometric validation may limit the interpretations. Though we have shown that phonemic awareness tasks differ in dyslexic children, the cross-sectional nature of the study prevents us from ascertaining a causal relationship between phonemic awareness and reading skills. Since phonemic awareness improves with schooling, a longitudinal study with preschoolers will help in clarifying its causal role in dyslexia in alphasyllabary languages.

The present study demonstrates that phonemic awareness deficits observed in dyslexia in alphasyllabary languages like Malayalam are more prominent in tasks where lexical support is absent, like pseudoword blending. These deficits vary across task types and linguistic complexity. Phonemic awareness shows improvement with age in typically developing children, while the improvement is patchy in children with dyslexia. The findings suggest that phonemic awareness deficits are a core feature of dyslexia across languages, though orthographic and linguistic characteristics of the writing system shape their manifestation. The results are important in designing phoneme-level interventions even in syllable-based writing systems.

## Data Availability

All data produced in the present study are available upon reasonable request to the authors. The word list used in this study is in Malayalam and is not included in the preprint due to language constraints. It is available from the corresponding author upon reasonable request.

## Declaration of Conflicting Interests

The authors declared no potential conflicts of interest with respect to the research, authorship, and/or publication of this article.

## Funding

The authors received no financial support for the research, authorship, and/or publication of this article.

## Notes

### Competing Interest Statement

The authors have declared no competing interest.

### Funding Statement

This study did not receive any funding

### Author Declarations

The study was approved by the institutional ethics committee of the Institute of Mental Health and Neurosciences, Kozhikode, India IMHANS/IEC/CPT/2024/006

